# Racial/Ethnic Differences in COVID-19-Traumatic Symptoms, Sleep, Coping Outcomes in a Group of New-Yorkers

**DOI:** 10.1101/2023.07.31.23293452

**Authors:** Judite Blanc, Azizi Seixas, Sean Small, Evan Auguste, Anthony Briggs, Yoann Sophie Antoine, Omonigho Michael Bubu, Girardin Jean-Louis

## Abstract

**Background:** Little research has examined within/between group predictors and mediators of race/ethnic differences or disparities in mental and sleep health outcomes arising from the experience of the COVID-19 pandemic.

**Objectives:** This study sought to evaluate the effect of COVID-19 experiences on trauma-related symptoms and sleep quality among a multiracial/ethnic sample in New York.

**Method:** This is a cross-sectional study conducted online among multiethnic adults (n=541) who experienced the pandemic in New York from September to November 2020. Comparisons of characteristics and mean scores by race/ethnicity status were conducted using one-way ANOVA and independent samples t-tests for continuous variables and chi-square tests for categorical variables. Multilinear regression was used for associations between social determinants of health and/or SES, trauma-related symptoms, coping, and sleep.

**Results:** Compared to Whites [Mean (SD)= (24.1(7.6)] and other group [Mean (SD)=24.9(8.2), Blacks [Mean (SD)=(26.3(6.4)] and Hispanics [Mean(SD)=(27.2(8.2)] reported higher level of peritraumatic distress [df= 3; F=4273; p=0.005). The prevalence of clinically significant PTSD symptoms was 21.4%(n=113): [Whites=31(16.3%); Blacks=28(25.7%); Hispanics=24(25%); and other groups=30(22.4%); x2 =4.93; p=0.177]. This rate doubled [48.3%(257)] when it comes to the overall clinically significant depression level. Compared to all subcategories, [Blacks=52(47.7%); Hispanics =62(64.6%); other group=66(49.3%)], depression symptoms were lower among Whites [77(39.9%; *x*2 =15.71; *p*=0.001]. We found a prevalence of insufficient sleep <6 hours of 41%(198): [Whites=69(39.4%); Blacks=43(41.7%); Hispanics=46(52.3%); other groups=40(34.2%); *x*2=12.21; *p*=0.057]. Several unique demographic predictors of PTSD emerged for distinct racial/ethnic groups. Among Blacks, sex [β = −0.22; p < .01] and employment [β = −0.159; p < .05] emerged as significant predictors for PTSD, but for no other racial/ethnic group. Interestingly, among Hispanics [β = −0.144; p = .064] and Blacks [β = −0.174; p = .0.076], coping strategies did not mitigate PTSD or depressive symptoms.

**Conclusion:** As New York and the rest of the world are trying to bounce back from the COVID-19 consequences, mental health outcomes are devastating, particularly among historically marginalized communities. This study provides insight into the emergency for policymakers to invest in racial justice programs and provide free access to culturally responsive mental health care for the most vulnerable groups.

## Introduction

COVID-19 remains an ongoing public health crisis with mental health implications. As an unprecedented and pervasive disaster, COVID-19 is characteristic of a potentially traumatic event defined by the DSM-5, posing a fear-inducing, existential threat, as evidenced by its lethality [1]. Additionally, COVID-19 and previous pandemics (e.g., SARS) feature additional stressors such as quarantine and lockdown measures, the confluence of which resulting in acute and long-term (post-)pandemic psychological impacts [2, 3]. New York was the epicenter of the coronavirus crisis in the United States for more than a month. As such, COVID-19 had devastating effects on the public health, economic, and personal structures upholding New York State. Federal data show that New York was one of the states most devastated by the pandemic, especially during the Spring and Summer months of 2020, with over 200,000 laboratory-confirmed infection cases, 20,000 COVID-19-related deaths, and over 1.7 million filings for unemployment by the end of April 2020 [4, 5]. Mental health effects of COVID-19 were visible in New York City, with increased calls to *NYC Well* (Mental Health Support) related to anxiety in March and April 2020, compared to January and February [6]. Following quarantine and lockdown orders, the internet searches of New Yorkers (assessed via Google Trends) after March 22, 2020, revealed a similar increase in symptoms. Specifically, searches for anxiety (18%), panic attack (56%), and insomnia (21%) rose significantly, whereas depression and suicide did not-suggesting the need for focused treatment during quarantine for New Yorkers [7]. Using the electronic health records of 2,358,318 patients from the New York City metropolitan region to examine changes in clinic psychiatric diagnoses between March 2020 and August 2021, Xiao et al. [8] found evidence further underscoring the severity of the psychological impact COVID-19 had on one of the epicenters of the US early in the pandemic. For example, they found that between February and August 2021, among patients without recent clinical psychiatric diagnoses, new psychosis, anxiety disorders, and mood disorders increased among both COVID-19-positive and negative persons (8). The greatest increases in psychiatric diagnoses among COVID-19-positive patients who were not hospitalized were anxiety (378.7%) and mood disorders (269.0%) [8]. Psychological consequences during COVID-19 were also higher among vulnerable populations such as under-resourced college students, healthcare workers, pregnant women, and caregivers in New York [9-12]. One study found the rates of insomnia symptoms of at least moderate severity in healthcare workers (HCWs) in New York City during the first wave to be 72.6% [13]. Another found rates of COVID-19-related acute stress, depression, and anxiety symptoms common among HCWs in New York at 57%, 48%, and 33%, respectively [14].

Moreover, it was revealed that the pandemic disproportionately affected Hispanics and Black New Yorkers. The COVID-19 literature describes racial/ethnic disparities in COVID-19 infections, hospitalizations, ICU admissions, and deaths, with Black and Hispanic people bearing the highest burden [15-19]. Similarly, racial/ethnic disparities in the harmful psychological consequences of COVID-19 have been demonstrated outside of New York [20, 21]. McKnight-Eily et al. [20] found disproportionately higher prevalence rates of self-reported suicidal ideation, depression, and increased or newly initiated substance abuse among Hispanic adults during the pandemic compared to Whites and Blacks [20]. Additionally, using two nationally representative US surveys with large sample sizes covering 2019 and 2020-21, Thomeer et al. [21] found that the mental health of racial minorities (Black and Hispanic) worsened disproportionately relative to Whites during the pandemic compared to before the pandemic, with significant increases in anxiety and depression. Specifically, in 2019, Blacks and Whites had rates of depression/anxiety at around 11% and Hispanics at 9.43%. And in 2020-21, Blacks (41.69%) and Hispanics (44.23%) had larger increases than Whites (34.31%) [21]. Additionally, they found that their findings suggested that the relative mental health advantage experienced in minoritized groups before the pandemic reversed during the pandemic [21]. The surprising results of Silverman et al. [11] in relation to the literature regarding racial disparities in negative psychological consequences reveal the complexities of examining social determinants of health and mental health. It is, thus, important to further study these disparities while considering the literature on social determinants of health. In the US, social determinants of negative health and lower socioeconomic status are inextricable from minoritizations and racial/ethnic minority status [28]. Racial and ethnic minority groups are more likely to live in crowded conditions, have less access to stable housing and livable wages, and have jobs that cannot shift remotely [10, 28]. Racial disparities have been observed in the interaction between economic stressors (e.g., food insecurity, job loss) and mental health consequences during COVID-19 [28-30]. As of April 12, 2020, near the height of the first wave in New York City, Hispanic and Black residents reported more housing-related stressors, such as fear of eviction and inability to pay rent [10], similarly reflected by disparities in national unemployment rates the same month [28]. The loss of health insurance and food insecurity/insufficiency were also more likely to be reported by Black and/or Hispanic populations [10, 30]. Of importance is that although these stressors increase during disasters such as COVID-19, the negative effect of this is distinctly worsened in groups that have already experienced socioeconomic difficulties [10, 28, 29]. Thus, pandemic-related, and concomitant socioeconomic stressors have implications for psychological distress during the pandemic, particularly for marginalized populations in New York.

With the pandemic being an ongoing, potentially traumatic event, it is valuable to gain insight into the mental health effects that precipitated early in the onset of COVID-19, especially in minoritized populations such as Blacks in New York. Disaster research literature among populations exposed to natural and man-made crises report a set of factors that may influence survivors’ resilience to deleterious disaster outcomes. These factors are usually regrouped into chronological order. In the case of COVID-19, we are referring to 1) pre-COVID-19 factors, 2) Peri-COVID-19 and lockdown factors, and 3) Post-COVID-19 lockdown factors (*Figure 1*) [9]. It is likely that mental health outcomes of the experience of the COVID-19 pandemic among New Yorkers are related to risk and protective factors at various socio-ecological levels, including individual (e.g., age, gender, and occupation), interpersonal (e.g., social support and/or coping strategies), institutional (e.g., public health policies, state regulations), and community (e.g., stigma, ethnicity, and cultural beliefs) factors. Indeed, Ghassabian et al. [12] found that among pregnant women and/or mothers of young children in a diverse NYC cohort, a history of depression (pre-COVID-19 factor), current financial difficulties, and being infected with COVID-19 (both peri-COVID-19 factors) were robustly associated with higher perceived stress. Thus, mental health is best understood as a dynamic state in which multiple intersecting social and environmental factors influence individual psychosocial development. In the womb, the mother’s mental and physical health status impacts the developing fetus [11]. Parent bonding and the home environment greatly affect the very early years. Thereafter, factors such as unemployment, poverty, physical health problems in adulthood, and levels of social and community connectedness in later life all have a part to play in influencing an individual’s ability to cope in disaster contexts. Adverse experiences can occur widely and cluster in some families, communities, and work settings already experiencing other destabilizing factors. Thus, one’s zip code plays a more prominent role than one’s genetic code in many ways, and the term “exposome” has been proposed as a new paradigm encompassing all aspects of human environmental exposures (not only genetic) starting from conception, complementing the genome and affecting health in a whole new way. It is, therefore, crucial that in disaster settings, if mental health problems are to be prevented, the social, economic, health, and ecological environment in which they are developing needs to be addressed [31].

**Figure 1:**
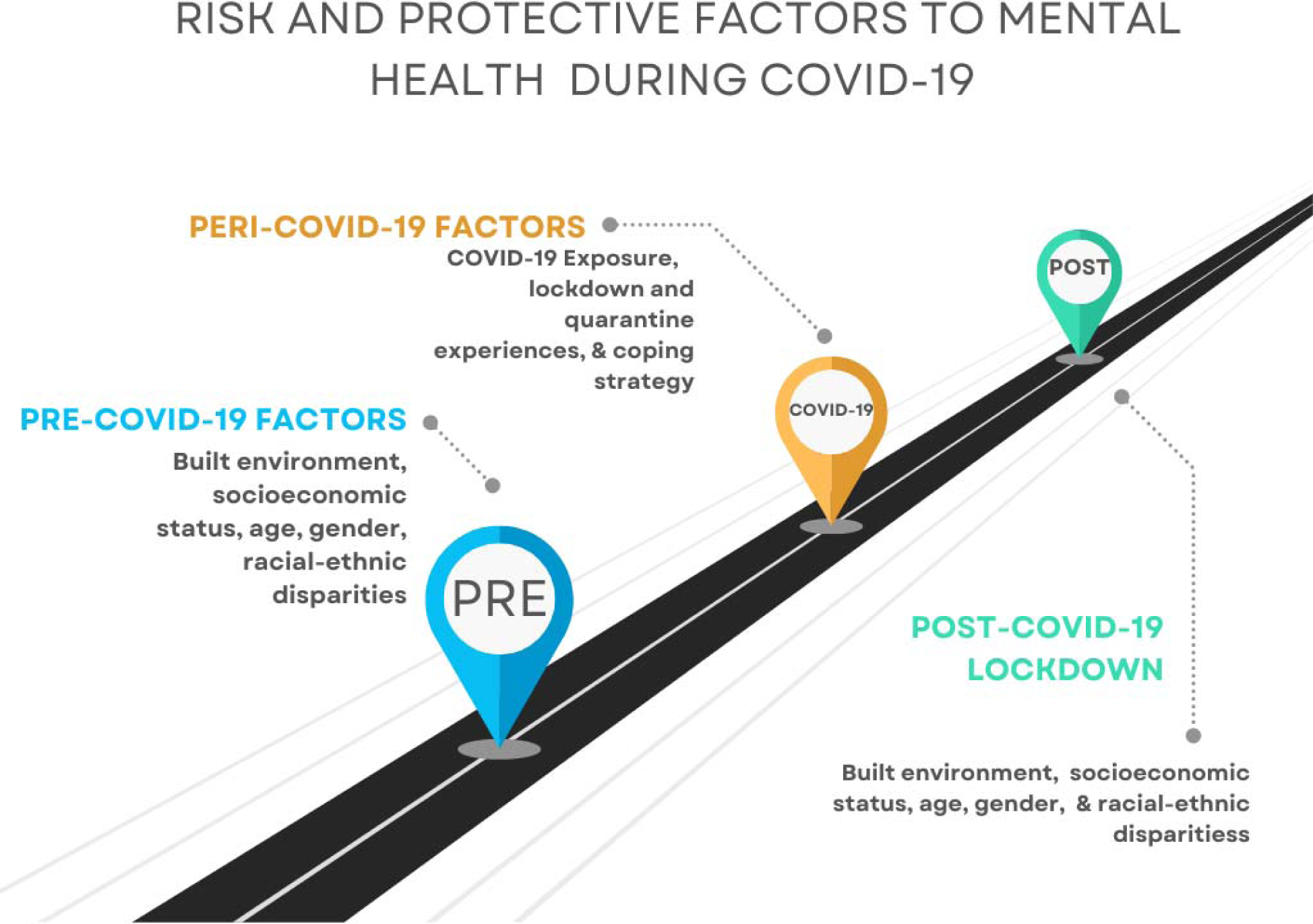
Pre, Peri, and Post-Covid-19 Lockdown Risk and Protective Factors For Mental Health

Hence, understanding specific pre-(built environment, socioeconomic status-SES, age, sex/gender, racial-ethnic disparities), peri-(serostatus, COVID-19 related experiences, the overall approach to lockdowns such as coping strategy), and post-lockdown (built environment, SES, age, sex/gender, racial-ethnic disparities) factors associated to trauma-related symptoms may inform the development of behavioral, translational interventions and programs to reduce the adverse mental health outcomes of the pandemic in vulnerable populations [32]. A few studies have examined within and between-group predictors and mediators of race/ethnic differences in trauma-related symptoms and sleep health outcomes during the US-COVID-19 pandemic. Aiming to fill this gap, we investigate the following research questions: 1) Among adult survivors of the COVID-19 pandemic during Spring 2020 in New York, what is the prevalence of trauma-related (PTSD and depression) and lower sleep quality symptoms? 2) Among these survivors in New York, what are the effects of socio-demographic factors and social determinants of health on trauma-related symptoms (PTSD and depression) and sleep health, and whether this relationship is mediated/moderated by race/ethnicity?

### Hypotheses

We hypothesized that:

1. Given the high fatality rate of the pandemic in New York City during Spring 2020 among Blacks and Hispanics, the prevalence of trauma-related symptoms such as PTSD, depression, and sleep problems would be highest among these racial/ethnic groups.
2. Social determinants of health and/or socioeconomic statuses (SES) factors such as sex, employment status, income, education, and marital status would explain within and between racial/ethnic group differences in trauma-related symptoms such as PTSD, depression, and sleep health;
3. Within racial/ethnic groups, peritraumatic distress will be the strongest predictor of trauma-related symptoms and sleep health after controlling for social determinants of health and/or SES factors.

## Method

### Participants

This cross-sectional study was conducted among adults who experienced the COVID-19 pandemic in New York State during the last semester of 2020. Participants were invited through ResearchMatch, Twitter, LinkedIn, secured email, and word of mouth within our community-based network throughout New York. Inclusion criteria comprised: 1) “Age ≥ 18 and living in New York at the time of the study since the first case of COVID-19 was reported on March 1st of 2020, and 2) Able to read and speak English. Exclusion criteria comprised: 1) Significant cognitive impairment or apparent during screening, and 2) History or current diagnosis of schizophrenia or other psychotic disorders. Between September and November 2020, 764 participants completed the screening questionnaire. Among them, 223 were excluded for not meeting the inclusion criteria or for incomplete data, and 541 were included for final statistical analysis. The New York University School of Medicine Institutional Review Board approved the study protocol. All participants provided electronic informed consent before enrollment.

### Procedures

The online survey included questions about demographics, including age, marital status, education level, employment status, and job types, socioeconomic status, place of birth, amount of time in the US. Previous medical history and COVID-19-related experiences included assessing distress due to COVID-19-specific stressors, trauma-related, sleep, and trauma-coping self-efficacy.

While the surveys were electronically captured, our outreach was multimodal, and it was the only way we could reach out to individuals, given the COVID-19 restrictions. We also utilized evidence-based community engagement strategies such as contacting faith-based organizations and community-based organizations who contacted their constituents through the phone and SMS.

### Measures

#### Post-Traumatic Stress Disorder (PTSD) Symptoms

Symptoms of PTSD in this study were assessed using the Posttraumatic Stress Disorder Checklist 5 (PCL-5). The PCL-5 is a 20-item self-report measure that assesses the presence and severity of DSM-5 symptoms of PTSD. Respondents were asked to rate how much they were bothered by a list of problems in the past month because of COVID-19. Examples of items are: Repeated, disturbing, and unwanted memories of the stressful experience; 2) Repeated, disturbing dreams of the stressful experience; 3) Suddenly feeling or acting as if the stressful experience was happening again (as if you were back there reliving it); to 4) Feeling very upset when something reminded you of the stressful experience [33]. Reliable PCL-5 cutoff scores range between 31-33 for a PTSD diagnosis [34]. According to the National Center for PTSD “Using the PTSD Checklist for DSM-5 (PCL-5)” a score of 31-33 or higher suggests the patient may benefit from PTSD treatment. PTSD patients can either be referred to a PTSD specialty clinic or offered an evidence-based treatment such as Prolonged Exposure (PE), Cognitive Processing Therapy (CPT), or Eye Movement Desensitization and Reprocessing (EMDR). When a patient’s score is lower than 31-33, this may indicate that they are experiencing subthreshold symptoms of PTSD or do not meet the criteria for PTSD, and this should be considered when planning their treatment. This questionnaire demonstrated good internal consistency (Cronbach’s α = .93) in the current sample.

#### Depressive Symptoms

Symptoms of depression were assessed using the Patient Health Questionnaire (PHQ-9), a multipurpose module for screening, diagnosing, monitoring, and measuring the severity of depressive symptoms. Using a Likert-scale ranging from (1) *Not at all* to (3) *Nearly every day*, PHQ-9 was used to assess how often participants were experiencing depressive symptoms, such as 1) Little interest or pleasure in doing things; 2) Feeling down, depressed, or hopeless; 3) Trouble falling or staying asleep or sleeping too much. The overall scores range from 0-27, with a cutoff score of 9 and above, indicating mild to severe depressive symptoms [35]. The PHQ-9 has been shown to be unidimensional, reliable, and mostly free of dimensional items functioning across countries during the pandemic [36]. This questionnaire demonstrated good internal consistency (Cronbach’s α = .89) in the current sample.

#### Sleep Quality

Sleep quality during the past month due to experiencing the COVID-19 pandemic was assessed using the Pittsburgh Sleep Quality Index (PSQI), which consists of 4 open-ended and 14 questions (18 questions in total) answered using event frequency and semantic scales. Examples of items include: “1) During the past month, what time have you usually gone to bed at night; 2) During the past month, how long (in minutes) has it usually takes you to fall asleep each night; 3) During the past month, how often have you had trouble sleeping because you cannot get to sleep within 30 minutes; 4) During the past month, how would you rate your sleep quality overall?” [37]. A cut-off score of 5 on the global scale was used to assess sleep quality among respondents [38]. We defined insufficient sleep as total sleep time ≤6 hours. In this present sample the seven component scores of the PSQI had a low level of internal reliability (Cronbach’s α = .53) compared to the high level of 0.83 reported by the validation study [37, 38].

#### COVID-19-Related Experiences and Distress

COVID-19-related experiences operationalized whether participants had been recently diagnosed with COVID-19 and/or had a close relative or family member recently diagnosed with COVID-19 and quantified/characterized home quarantine experience. COVID-19-related experiences were assessed using the “*COVID-19-related experiences questionnaire*” (*Yes/No items)* developed by the authors. Home quarantine experience included 8 items about whether participants: 1) Wear a mask when in contact with family members; 2) Take their temperature twice a day; 3) Sleep in a separate room; or 4) Do not go to community activities (functions, shopping, etc.), among others. COVID-19 quarantine experience scores ranged from 1-8 in our study. This questionnaire demonstrated good internal consistency (Cronbach’s α = .72) in the current sample.

#### Peritraumatic Distress

The Peritraumatic Distress Inventory (PDI) consists of 13 self-reported items originally developed based on DSM-4 criteria for PTSD diagnosis [39]. It was used to assess the intensity of emotional and physiological distress that participants were experiencing during the pandemic. Using a 5-point Likert scale with the following anchors: 0 = Not at All True; 1 = Slightly True; 2 = Somewhat True; 3 = Very True; and 4 = Extremely True, participants were asked to rate feelings such as 1) Helplessness; 2) Sadness or grief, and 3) Feeling afraid for their safety. Total scores ranged from 0–to 52. The original English PDI version demonstrated that a cutoff score of 23 (sensitivity = 71%; specificity = 73%) is a good index to predict clinically elevated PTSD among traumatically injured patients after 30 days [40]. The PDI demonstrated good internal consistency (Cronbach’s α = .87) in the current sample.

#### Trauma Coping Self-Efficacy

The Trauma Coping Self-Efficacy (CSE-T) comprises 20 questions that assess participants’ perceived ability to cope with challenges and demands from a traumatic event similar to the COVID-19 pandemic. Respondents were asked to rate their capability to handle a series of traumatic pandemic demands on a 7-point scale from 1 (*not at all capable*) to 7 (*totally capable*), in dealing with the following questions: 1) Deal with my emotions (anger, sadness, depression, anxiety) since I experienced my trauma; 2) Deal with the impact the trauma has had on my life; 3) Get my life back to normal, or 4) Control distressing thoughts about the traumatic experience [41]. This measure demonstrated reliability in assessing coping self-efficacy in stress associated with the COVID-19 pandemic and good internal consistency (Cronbach’s α = .92) in the current sample [42].

#### Statistical Analysis

Participant characteristics are reported for the sample. Comparisons of participant characteristics based on race/ethnicity were conducted using one-way ANOVA and independent samples t-tests for continuous variables and chi-square tests for categorical variables. Characteristics were reported as means with standard deviations or as frequencies. Associations between social determinants of health and/or SES and trauma-related symptoms, sleep, and coping were examined using stratified multilinear regression analysis by race/ethnicity. The model examined the association between age, NYC residency, sex, employment status, peritraumatic distress and coping with trauma-related symptoms, and sleep health by race/ethnicity. The strength of the association between SES characteristics with trauma-related symptoms, sleep health, and coping was evaluated by interpreting the strength of the regression parameter *Beta* as well as the variance explained by the model (defined as the R^2^ adjusted).

## Results

### Participants Characteristics

The sample consisted of 541 participants recruited across New York State. The majority were female [373 (68.9%)], and the mean age was 40.9 years (SD=15). As indicated in Table 1, 36.6% of volunteers identified as White, 20.5% as Black, 17.9% as Hispanic, and 25% identified as either Asian (42), Native-American (1), Pacific-Islander (1), or Unknown (91) (classified as “Other”). Among our sample, 342(63.2%) had a college education [Whites= 116(58.9%); Blacks=81(73%); Hispanics= 66(68%); Other groups=79(59.4%); x2 =; p<0.001]. An amount of 123(25.2%) were essential workers, [Whites=50(26.2%); Blacks=29(30.5%); Hispanics=13(16.3%); other groups=31(25.4%); *x*2 =4.92; p=0.177.].

**Table 1:**
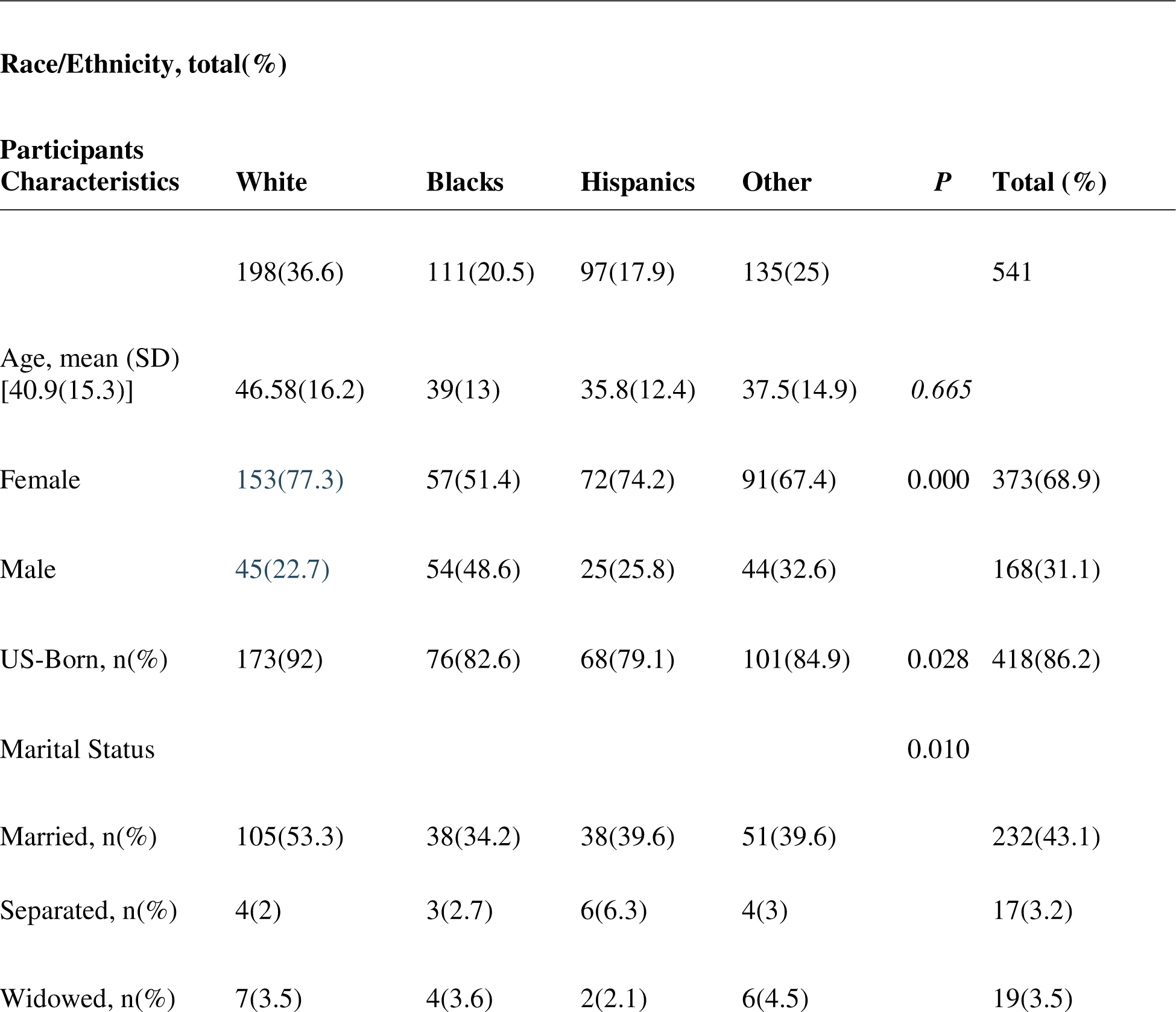
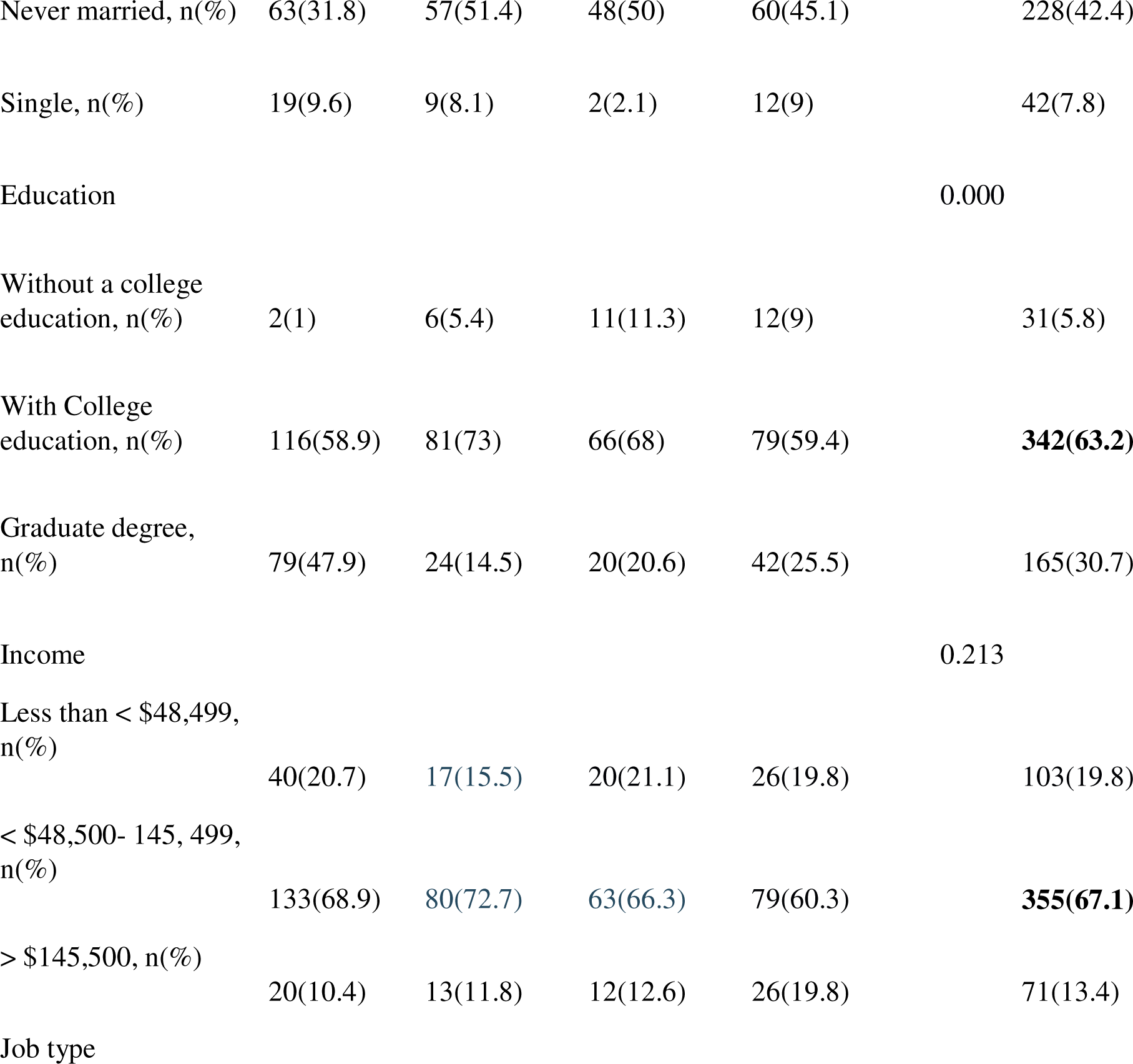
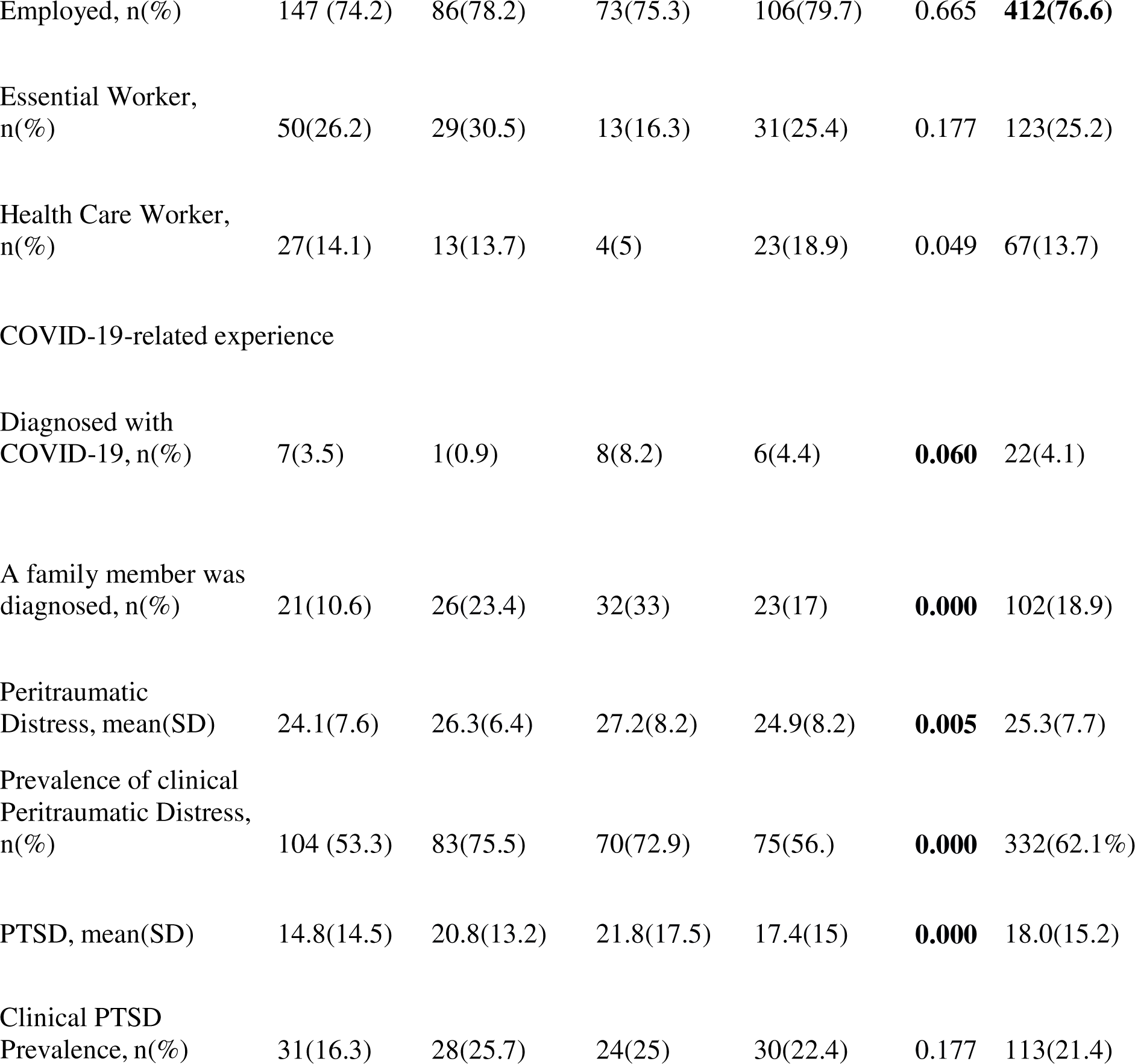
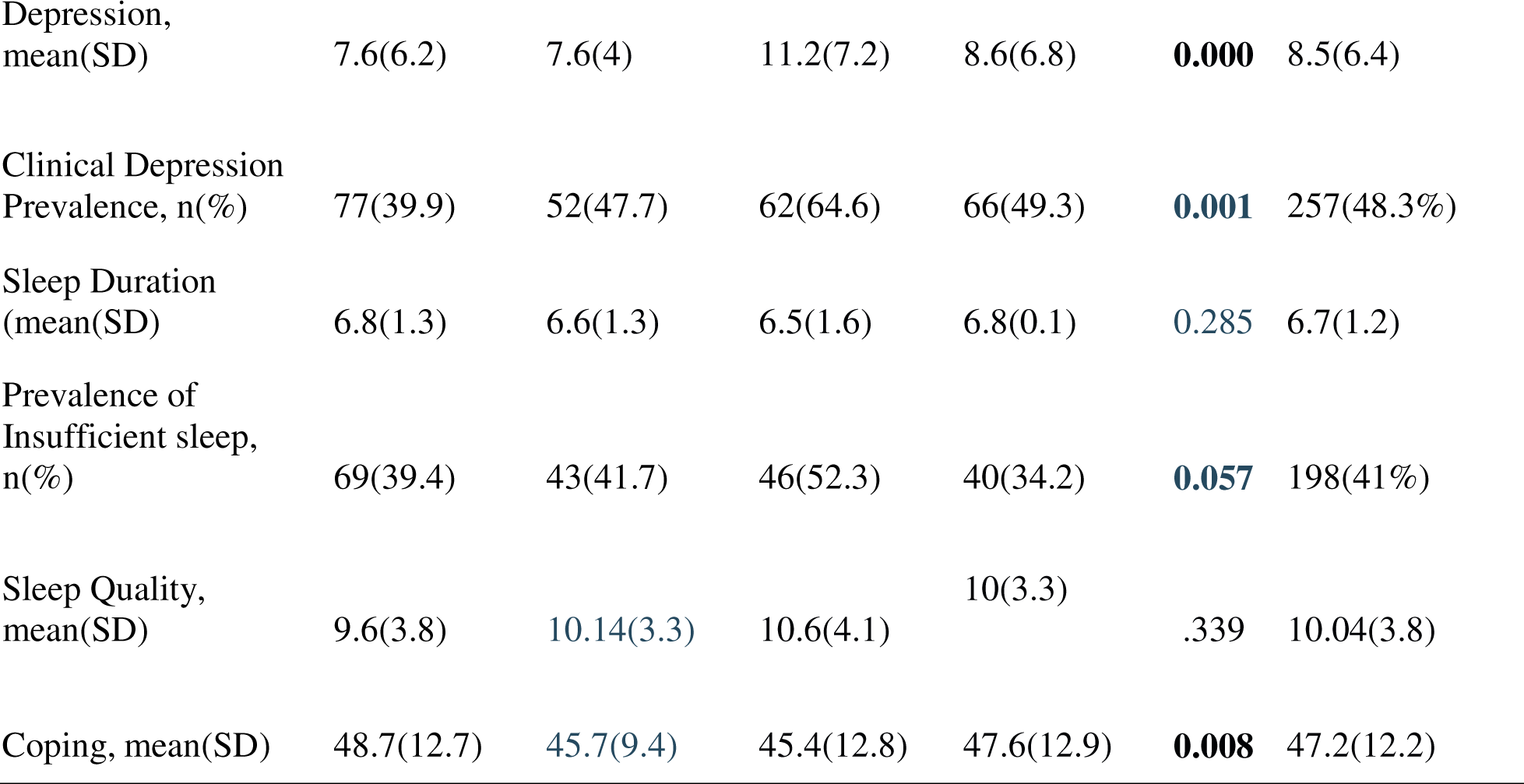
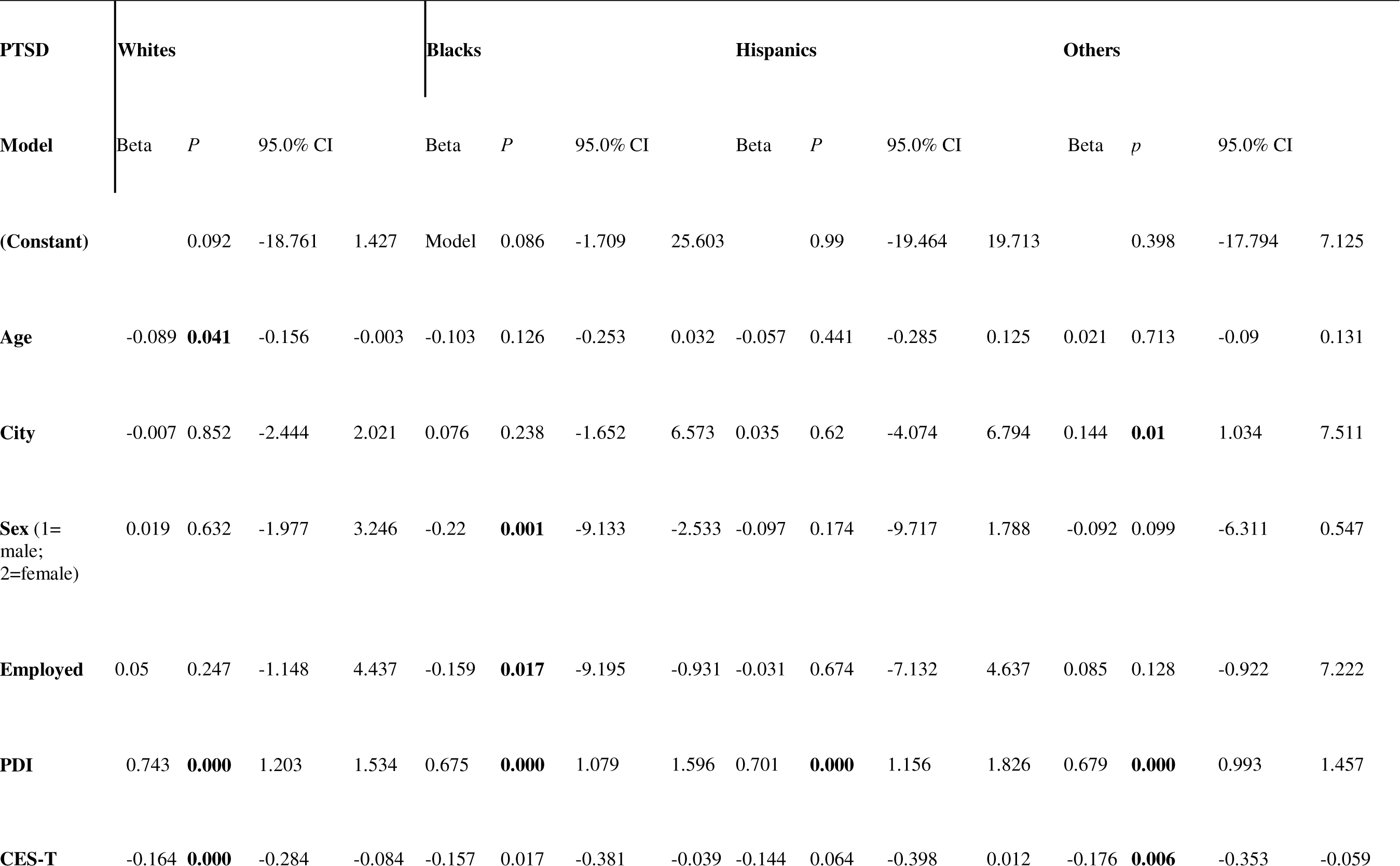
Characteristics of the Participants by Race/Ethnicity, N= 541, (** = P < 0.05) Race/Ethnicity, total(%)

### The Prevalence of Peritraumatic Distress, PTSD, Depression and Sleep Problems

As noted in Table 1, there were significant racial and ethnic differences/disparities in the prevalence of clinically significant peritraumatic distress, depressive, and PTSD symptoms. Concerning COVID-19 experiences, 4.1%(22) suffered from COVID-19 [Whites (7); Blacks (1); Hispanics (8); other groups(6); *x*^2^ =7.39; *p*=.06]. A total of 102(18.9%) had a family member who was diagnosed with COVID-19 [Whites=21(10.6%); Blacks=26(23.4%); Hispanics= 32(33%); Other groups=23(17%); *x*^2^ =23.27; *p*=0.000]. Compared to Whites [Mean(SD)=(24.1(7.6)] and th other groups [Mean(SD)=24.9(8.2), Blacks [Mean(SD)=(26.3(6.4)] and Hispanics [Mean(SD)=(27.2(8.2)] reported higher level of peritraumatic distress [df= 3; F=4273; *p*=0.005).

We found a prevalence of clinically significant PTSD symptoms of 21.4%(n=113) [Whites=31(16.3%); Blacks=28(25.7%); Hispanics=24(25%); and other groups=30(22.4%); x2 =4.93; *p*=0.177]. This rate doubled [48.3%(257)] when it comes to the overall clinically significant depression level; and compared to all subcategories [Blacks=52(47.7%); Hispanics =62(64.6%); other groups=66(49.3%); *x*^2^ =15.71; p=0.001], depression symptoms were lower among Whites[77(39.9%]. Finally, we found a prevalence of insufficient sleep <6 hours of 41%(198) [Whites=69(39.4%); Blacks=43(41.7%); Hispanics=46(52.3%); other groups=40(34.2%); *x*^2^ =12.21; *p*=0.057].

### The Moderating Role of Race/Ethnicity in the Relationship Between Demographics, Peritraumatic Distress, Coping, and Trauma-related Symptoms

Table 2 shows the results for this section. Regarding PTSD, peritraumatic distress emerged as the strongest independent predictor among all variables entered the model for White [β = 0.743; p < .001], Black [β = 0.675; p < .001], Hispanic [β = 0.701; p < .001], and other participants [β = 0.679; p < .001]. In addition, several demographic predictors of PTSD emerged for distinct racial/ethnic groups. Sex [β = −0.22; p < .01] and employment [β = −0.159; p < .05] emerged as significant predictors for PTSD for Black participants only. Age [β = −0.089; p < .05] emerged as a significant predictor of PTSD for White participants only. Interestingly, Trauma coping self-efficacy was a strong independent predictor of PTSD for all groups except Hispanic participants [β = −0.144; p = .064]. The final model accounted for 66% of the PTSD variance [R^2^ = 0.666; *p* < 0.001].

**Table 2:**
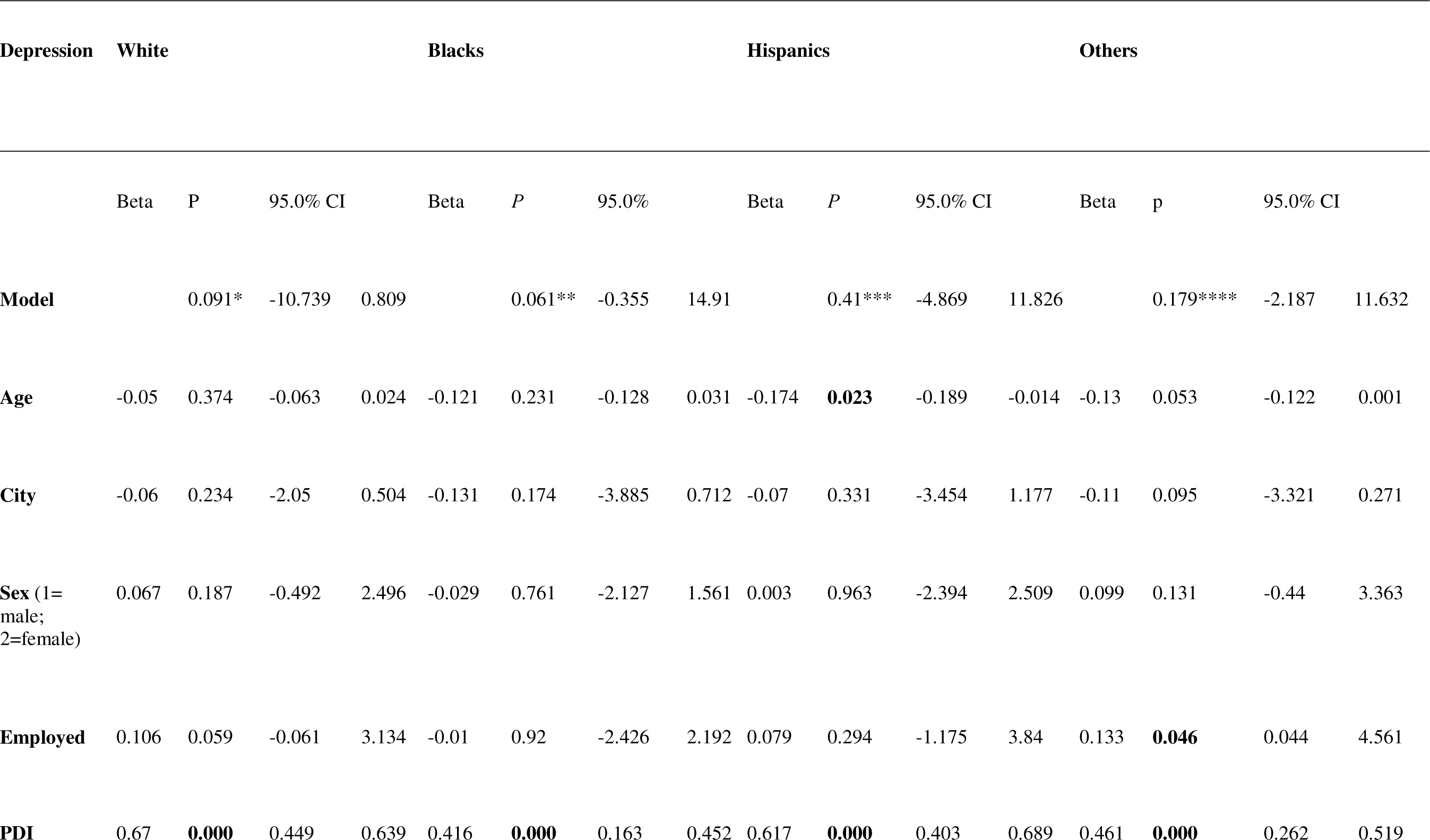
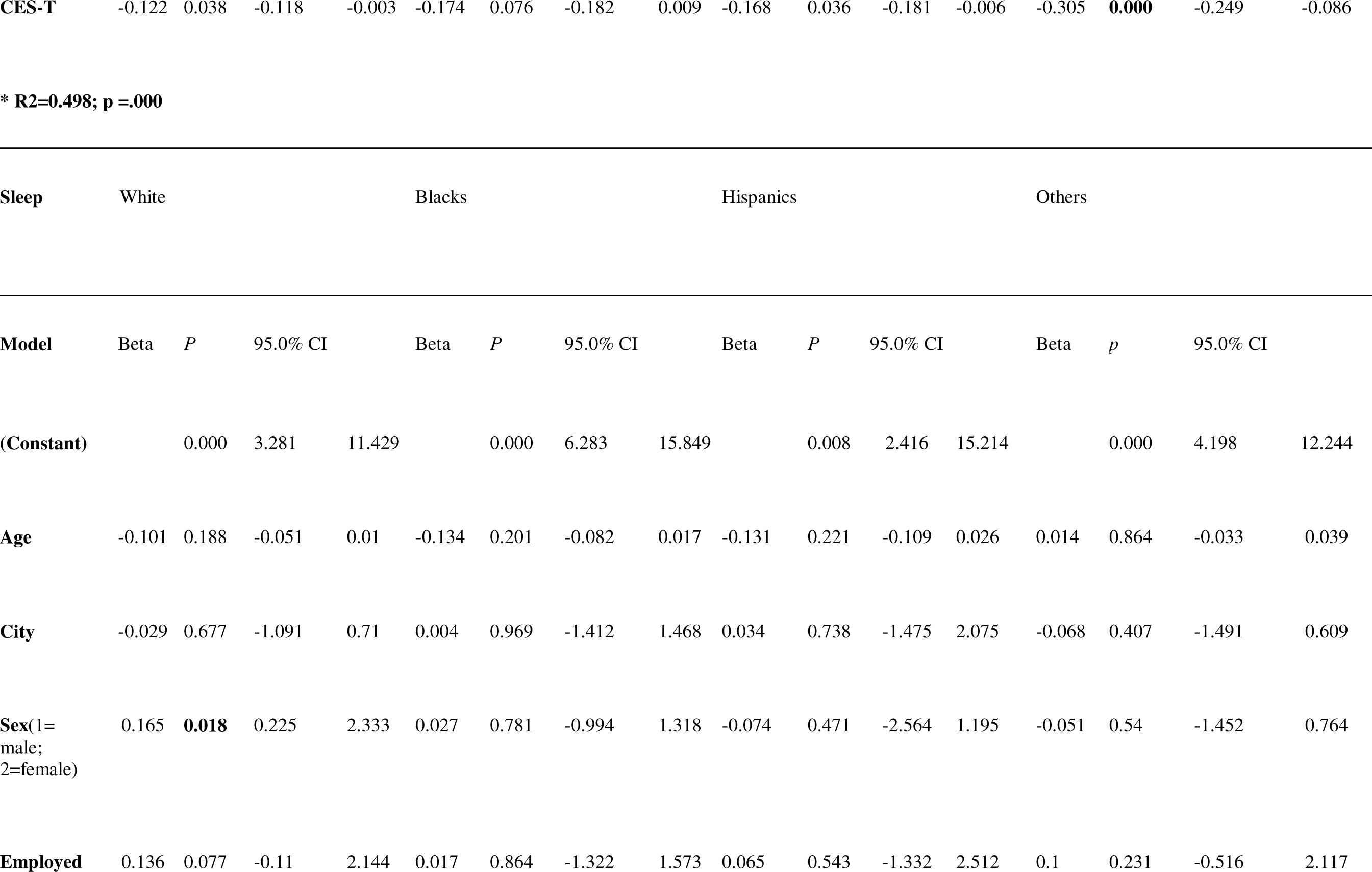
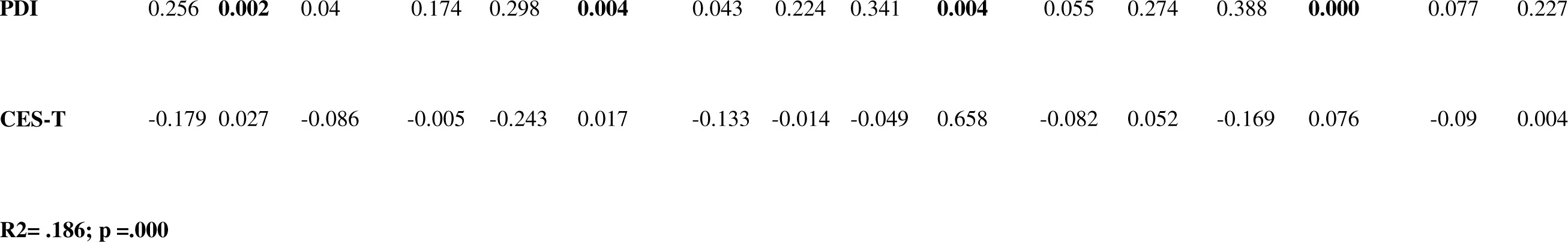
Stratified multilinear regression analyses predicting PTSD, Depression and Sleep Health by Race/Ethnicity.

Similarly, peritraumatic distress was the strongest independent predictor of depression for White [β = 0.670; p < .001], Black [β = 0.416; p < .001], Hispanic [β = 0.617; p < .001], and other participants [β = 0.461; p < .001]. Employment status uniquely predicted depression for participants categorized as Other [β = 0.133; p < .05]. Trauma-coping self-efficacy predicted depression in all groups except Black participants [β = −0.174; p = 0.076]. The final model accounted for significant variance in peritraumatic distress [R^2^ = 0.498; *p* < 0.001].

Finally, peritraumatic distress was again the strongest independent predictor for sleep health quality among White [β = 0.256; p < .01], Black [β = 0.298; p < .01], Hispanic [β = 0.341; p < .01], and other participants [β = 0.388; p < .001]. Sex was a unique predictor of sleep quality for White participants [β = 0.165; p < .05]. Trauma-coping self-efficacy was a significant predictor for White [β = −0.179; p < .05] and Black [β = −0.243; p < .05] participants. The final model accounted for 18% of the sleep health variance [R^2^ = 0.186; *p* < 0.001].

According to the multilinear regression results in table 3, when the entire sample is considered together and accounting for social determinants of health, peritraumatic distress and trauma coping strategies remain the most important predictors of PTSD [(β = .711; *p*= 0.000); (β = − 0.158; p= 0.000)], depression [(β = .565; *p*= 0.000); (β = −.191; p= 0.000)], and sleep quality [(β = 0.317; *p*= 0.000); (β = ^-0.152^; p= 0.001)], followed by geographical location in New York and sex. However, race/ethnicity is no longer a significant predictor of PTSD, depression, and sleep quality.

**Table 3:**
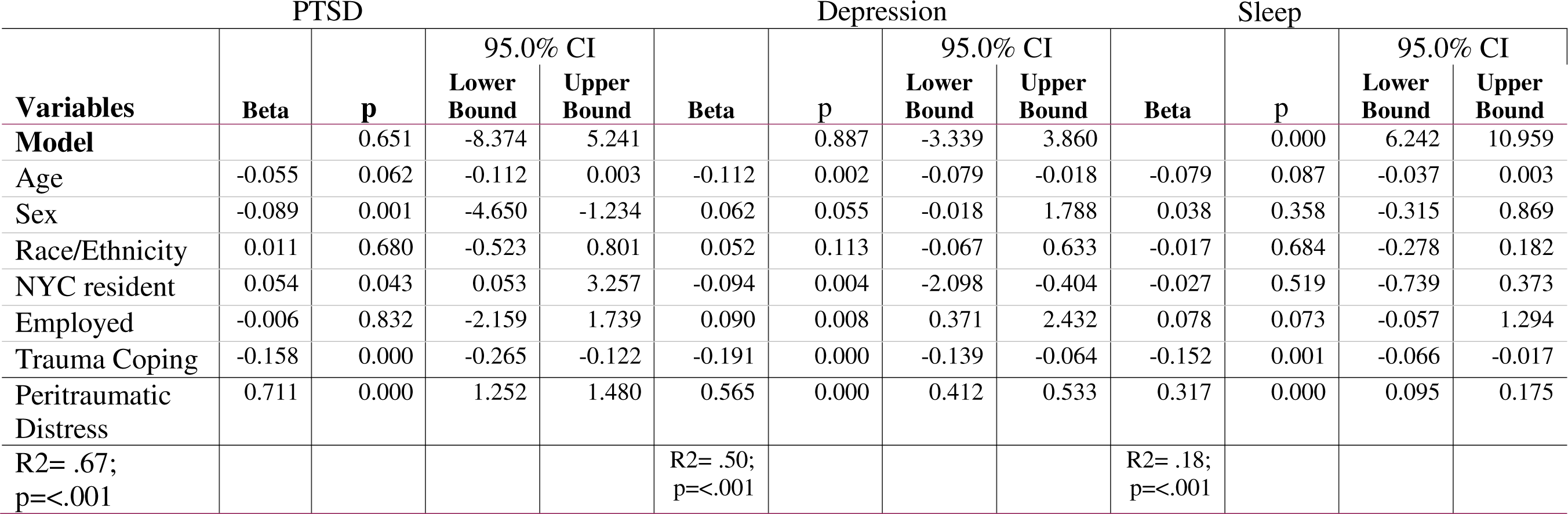
Multilinear regression analyses with social determinants of health, race/ethnicity and sex predicting PTSD, Depression and Sleep Health.

## Discussion

Among a predominantly female, college-educated, and multi-ethnic group of survivors of the COVID-19 pandemic in New York State, this study assessed: 1) the prevalence of trauma-related symptoms and sleep quality; 2) the effects of socio-demographic factors and social determinants of health on trauma-related symptoms and sleep health, and whether this relationship is mediated/moderated by race/ethnicity. We found widespread and significant racial/ethnic differences in COVID-19 trauma-related symptoms such as peritraumatic distress, PTSD, depression, and to a certain extent insufficient sleep.

Our first hypothesis regarding the effect of race/ethnicity was supported as it revealed that historically disadvantaged participants reported greater symptoms and levels of clinical peritraumatic distress, PTSD, and depression. For example, among our sample, Hispanics obtained the highest scores for peritraumatic distress, PTSD, and depression, followed by Black participants who also reported greater peritraumatic distress levels and PTSD compared to Whites and the other non-Hispanic group. This finding is consistent with research that has found that relative to other ethnic/racial groups, Whites are less likely to develop PTSD in response to traumatic exposure [45]. Previous research has suggested that, unlike their white counterparts, Blacks may be at higher risk for PTSD due to their lived experiences with historical, economic, and social trauma [46]. As such, COVID-19 seems to further exacerbate those social determinants of health among this group, contributing to the onset of PTSD symptoms. Some suggest that this stronger susceptibility to PTSD among Black people, specifically Black Americans, is due to increased exposure to racial discrimination that contributes to PTSD [47].

### Race, Sex, and Employment in PTSD

Studies show that job loss, loss of health insurance coverage, social isolation, and housing-related stressors increased the risk of PTSD and chronic psychological distress during COVID-19 [43, 44]. Our second hypothesis regarding the relevance of social determinants, e.g., employment status and male sex was partially supported as sex and employment emerged as significant predictors for PTSD for Black participants only [49]. This surprising result could be explained by the pre-existing socio-economic adversities faced by Black male New Yorkers and the fact that the prevalence of COVID-19 was higher among men. As of April 12, 2020, near the height of the first wave in New York City, Hispanic and Black residents reported more housing-related stressors, such as fear of eviction and inability to pay rent [10]. The loss of health insurance and social isolation were also more likely to be reported by Hispanic populations [10]. It is worth considering how the disproportionate loss from the pandemic for Black people was experienced in association with the Georges Floyd killing, and national uprisings against police brutality and may have contributed to elevated PTSD risk and severity [27] among Black males. Other research suggests that, even when considering race, social support is the strongest predictor for the trajectory of trauma symptoms [48]. This would mean that it is possible that disparate social disruption due to the pandemic impacted the severity and expression of trauma-related symptoms, as opposed to purely understanding pandemic-related stress as only race-related. This is consistent with the disproportionate economic stressors, including fears around eviction, that Black and Hispanic residents in NYC have reported [10].

### The Influence of Peritraumatic Distress During COVID-19

As expected, we found that across our sample, peritraumatic distress was the strongest predictor of trauma-related symptoms and sleep quality after controlling for social determinants or SES and coping skills. This is consistent with prior literature that has demonstrated COVID-19-related distress to be one of the strongest predictors of psychological problems like PTSD and insomnia [50, 51].

### Limitations

Although the significance of our findings through the fact that it is advancing data on the mental health consequences of the experiences of the COVID-19 pandemic from a racial/ethnic perspective in New York, it is worth noting some weaknesses that could impact the generalizability of the data. Data was gathered remotely with a convenient sampling method that could be a threat to the generalizability of our findings to the entire New York State population. Additionally, our findings are limited to self-reported instruments and there is no follow-up to assessment to evaluate the evolution of our study participants.

## Conclusion

In this study examining the COVID-19 trauma-related symptoms among a multiethnic group of New Yorkers, we found severe and widespread racial/ethnic differences/disparities in the level of peritraumatic distress, symptoms of PTSD, depression, and insufficient sleep. As New York and the rest of the world are trying to bounce back from the COVID-19 consequences, the mental health outcomes are devastating, particularly among historically marginalized communities. This study provides insight into the emergency for policymakers to invest in racial justice and healing initiatives across New York and nationally, and to provide free access to culturally responsive mental health care to tackle the racial/ethnic disparities in mental health.

## Data Availability

All data produced in the present study are available upon reasonable request to the authors.

https://www.ncbi.nlm.nih.gov/pmc/articles/PMC8135650/

## References

1. American Psychiatric Association. Diagnostic and Statistical Manual of Mental Disorders. 5th ed. Washington D.C.: 2013.

2. Veldhuis CB, Nesoff ED, McKowen ALW, et al: Addressing the critical need for long-term mental health data during the COVID-19 pandemic: Changes in mental health from April to September 2020. Prev Med 2021; 146:106465. DOI: 10.1016/j.ypmed.2021.106465

3. Brooks SK, Webster RK, Smith LE, et al. The psychological impact of quarantine and how to reduce it: rapid review of the evidence. Lancet. 2020;395(10227):912–920. doi:10.1016/S0140-6736(20)30460-8

4. Gharehgozli O, Nayebvali P, Gharehgozli A, Zamanian Z: Impact of COVID-19 on the Economic Output of the US Outbreak’s. Econ Disaster Clim Chang 2020; 1–13. DOI:10.1007/s41885-020-00069-w

5. Yang W, Kandula S, Huynh M, et al: Estimating the infection-fatality risk of SARS-CoV-2 in New York City during the spring 2020 pandemic wave: a model-based analysis [published correction appears in Lancet Infect Dis. 2021 Jan;21(1):e1]. Lancet Infect Dis 2021; 21(2):203–212. DOI:10.1016/S1473-3099(20)30769-6

6. Magas, I., Norman, C., Baxter, A., Harrison, M. Impact of COVID-19 on Mental Health in New York City. New York City Department of Health and Mental Hygiene, September 2020. https://www1.nyc.gov/site/doh/about/press/pr2020/new-data-on-covid-19-mental-health.page

7. Stijelja S, Mishara BL. COVID-19 and Psychological Distress-Changes in Internet Searches for Mental Health Issues in New York During the Pandemic. JAMA Intern Med. 2020;180(12):1703–1706. doi:10.1001/jamainternmed.2020.3271

8. Xiao Y, Sharma MM, Thiruvalluru RK, et al. Trends in psychiatric diagnoses by COVID-19 infection and hospitalization among patients with and without recent clinical psychiatric diagnoses in New York city from March 2020 to August 2021. Transl Psychiatry. 2022;12(1):492. Published 2022 Nov 21. doi:10.1038/s41398-022-02255-8

9. Blanc J, Briggs AQ, Seixas AA, et al: Addressing psychological resilience during the coronavirus disease 2019 pandemic: a rapid review. Curr Opin Psychiatry 2021; 34(1):29–35 DOI: 10.1097/YCO.0000000000000665

10. Rudenstine S, McNeal K, Schulder T, Ettman CK, Hernandez M, Gvozdieva K, & Galea S: Depression and Anxiety During the COVID-19 Pandemic in an Urban, Low-Income Public University Sample. J Trauma Stress 2021; 34:12–22. DOI: 10.1002/jts.22600

11. Silverman ME, Burgos L, Rodriguez ZI, et al. Postpartum mood among universally screened high and low socioeconomic status patients during COVID-19 social restrictions in New York City. Sci Rep. 2020;10(1):22380. Published 2020 Dec 24. doi:10.1038/s41598-020-79564-9

12. Ghassabian A, Jacobson MH, Kahn LG, Brubaker SG, Mehta-Lee SS, Trasande L. Maternal Perceived Stress During the COVID-19 Pandemic: Pre-Existing Risk Factors and Concurrent Correlates in New York City Women. Int J Public Health. 2022;67:1604497. Published 2022 Apr 11. doi:10.3389/ijph.2022.1604497

13. Abdalla M, Chiuzan C, Shang Y, et al: Factors Associated with Insomnia Symptoms in a Longitudinal Study among New York City Healthcare Workers during the COVID-19 Pandemic. Int J Environ Res Public Health. 2021; 18(17):8970. DOI: 10.3390/ijerph18178970

14. Shechter A, Diaz F, Moise N, et al: Psychological distress, coping behaviors, and preferences for support among New York healthcare workers during the COVID-19 pandemic. Gen Hosp Psychiatry 2020; 66:1–8. DOI: 10.1016/j.genhosppsych.2020.06.007

15. Behbahani S, Smith CA, Carvalho M, et al: Vulnerable Immigrant Populations in the New York Metropolitan Area and COVID-19: Lessons Learned in the Epicenter of the Crisis. Acad Med 2020; 95(12):1827–1830. DOI: 10.1097/ACM.0000000000003518

16. Thompson CN, Baumgartner J, Pichardo C, et al: COVID-19 Outbreak - New York City, February 29-June 1, 2020 [published correction appears in MMWR Morb Mortal Wkly Rep. 2020 Dec 18;69(50):1930]. MMWR Morb Mortal Wkly Rep. 2020; 69(46):1725–1729. DOI:10.15585/mmwr.mm6946a2

17. Renelus BD, Khoury NC, Chandrasekaran K, et al: Racial Disparities in COVID-19 Hospitalization and In-hospital Mortality at the Height of the New York City Pandemic. J Racial Ethn Health Disparities 2020; 1–7. DOI:10.1007/s40615-020-00872-x

18. Ogedegbe G, Ravenell J, Adhikari S, et al: Assessment of Racial/Ethnic Disparities in Hospitalization and Mortality in Patients With COVID-19 in New York City. JAMA Netw Open 2020; 3(12):e2026881. DOI: 10.1001/jamanetworkopen.2020.26881

19. Holtgrave DR, Barranco MA, Tesoriero JM, et al: Assessing racial and ethnic disparities using a COVID-19 outcomes continuum for New York State. Ann Epidemiol 2020; 48:9–14. DOI: 10.1016/j.annepidem.2020.06.010

20. McKnight-Eily LR, Okoro CA, Strine TW, et al. Racial and Ethnic Disparities in the Prevalence of Stress and Worry, Mental Health Conditions, and Increased Substance Use Among Adults During the COVID-19 Pandemic - United States, April and May 2020. MMWR Morb Mortal Wkly Rep. 2021;70(5):162–166. Published 2021 Feb 5. doi:10.15585/mmwr.mm7005a3

21. Thomeer MB, Moody MD, Yahirun J. Racial and Ethnic Disparities in Mental Health and Mental Health Care During The COVID-19 Pandemic [published online ahead of print, 2022 Mar 22]. J Racial Ethn Health Disparities. 2022;1–16. doi:10.1007/s40615-022-01284-9

22. Gur RE, White LK, Waller R, et al. The Disproportionate Burden of the COVID-19 Pandemic Among Pregnant Black Women. Psychiatry Res. 2020;293:113475. doi:10.1016/j.psychres.2020.113475

23. Park J. Who is hardest hit by a pandemic? Racial disparities in COVID-19 hardship in the U.S., International Journal of Urban Sciences. 2021. 25:2, 149–177, DOI: 10.1080/12265934.2021.1877566

24. Garcia MA, Homan PA, García C, Brown TH. The Color of COVID-19: Structural Racism and the Disproportionate Impact of the Pandemic on Older Black and Latinx Adults. The Journals of gerontology. Series B, Psychological Sciences and Social Sciences. 2021 Feb;76(3):e75–e80. DOI: 10.1093/geronb/gbaa114.

25. Macias Gil R, Marcelin JR, Zuniga-Blanco B, et al. COVID-19 Pandemic: Disparate Health Impact on the Hispanic/Latinx Population in the United States. The Journal of Infectious Diseases. 2020 Oct;222(10):1592–1595. DOI: 10.1093/infdis/jiaa474.

26. Laurencin CT, Walker JM. A Pandemic on a Pandemic: Racism and COVID-19 in Blacks. Cell Systems. 2020 Jul;11(1):9–10. DOI: 10.1016/j.cels.2020.07.002. PMID: 32702320; PMCID: PMC7375320.

27. Airhihenbuwa, CO: From 1619 to COVID-19: A Double Pandemic. Health Promot Pract 2020; 21(6):857–858. DOI: 10.1177/1524839920953819

28. Lopez L, Hart LH, Katz MH. Racial and Ethnic Health Disparities Related to COVID-19. JAMA. 2021;325(8):719–720. doi:10.1001/jama.2020.26443

29. Santos, GM., Ackerman, B., Rao, A. et al. Economic, Mental Health, HIV Prevention and HIV Treatment Impacts of COVID-19 and the COVID-19 Response on a Global Sample of Cisgender Gay Men and Other Men Who Have Sex with Men. AIDS Behav 25, 311–321 (2021). DOI: 10.1007/s10461-020-02969-0

30. Nagata, J. M., Ganson, K. T., Whittle, H. J., Chu, J., Harris, O. O., Tsai, A. C., & Weiser, S. D. (2021). Food Insufficiency and Mental Health in the U.S. During the COVID-19 Pandemic. *American* Journal of Preventive Medicine, 60(4), 453–461. DOI: 10.1016/j.amepre.2020.12.004

31. Kousoulis AA, Goldie I. A Visualization of a Socio-Ecological Model for Urban Public Mental Health Approaches. Front Public Health. 2021;9:654011. Published 2021 May 13. doi:10.3389/fpubh.2021.654011

32. WEISÆTH, L. Vulnerability and protective factors for posttraumatic stress disorder. Psychiatry and Clinical Neurosciences 1998; 52: S39–S44. https://doi.org/10.1046/j.1440-819.1998.0520s5083.x

33. Blevins CA, Weathers FW, Davis MT, et al: The Posttraumatic Stress Disorder Checklist for DSM-5 (PCL-5): Development and Initial Psychometric Evaluation. J Trauma Stress 2015; 28(6):489–498. DOI: 10.1002/jts.22059

34. Bovin MJ, Marx BP, Weathers FW, Gallagher MW, Rodriguez P, Schnurr PP, & Keane TM: Psychometric properties of the PTSD Checklist for Diagnostic and Statistical Manual of Mental Disorders-Fifth Edition (PCL-5) in Veterans. Psychological Assessment 2016;28(11), 1379–1391. DOI: 10.1037/pas0000254

35. Seo JG & Park SP: Validation of the Patient Health Questionnaire-9 (PHQ-9) and PHQ-2 in patients with migraine. J Headache Pain 2015; 16:65. DOI:10.1186/s10194-015-0552-2

36. Shevlin M, Butter S, McBride O, et al. Measurement invariance of the Patient Health Questionnaire (PHQ-9) and Generalized Anxiety Disorder scale (GAD-7) across four European countries during the COVID-19 pandemic. BMC Psychiatry. 2022;22(1):154. Published 2022 Mar 1. doi:10.1186/s12888-022-03787-5

37. Buysse DJ, Reynolds CF 3rd, Monk TH, et al: The Pittsburgh Sleep Quality Index: a new instrument for psychiatric practice and research. Psychiatry Res 1989; 28(2):193–213. DOI: 10.1016/0165-1781(89)90047-4

38. Pilz LK, Keller LK, Lenssen D, & Roenneberg T: Time to rethink sleep quality: PSQI scores reflect sleep quality on workdays. Sleep 2018; 41(5). DOI: 10.1093/sleep/zsy029

39. Brunet A, Weiss DS, Metzler TJ, et al: The Peritraumatic Distress Inventory: a proposed measure of PTSD criterion A2. Am J Psychiatry 2001; 158(9):1480–1485. DOI: 10.1176/appi.ajp.158.9.1480

40. Bunnell BE, Davidson TM, & Ruggiero KJ: The Peritraumatic Distress Inventory: Factor structure and predictive validity in traumatically injured patients admitted through a Level I trauma center. J Anxiety Disord 2018; 55:8–13. DOI: 10.1016/j.janxdis.2018.03.002

41. Benight CC, Shoji K, James LE, et al: Trauma Coping Self-Efficacy: A Context-Specific Self-Efficacy Measure for Traumatic Stress. Psychol Trauma 2020; 7(6):591–599. DOI: 10.1037/tra0000045

42. Shahrour G & Dardas LA: Acute stress disorder, coping self-efficacy and subsequent psychological distress among nurses amid COVID-19. J Nurse Manag 2020; 28(7):1686–1695. DOI: 10.1111/jonm.13124

43. Asmundson G, Paluszek MM, Landry CA, et al: Do pre-existing anxiety-related and mood disorders differentially impact COVID-19 stress responses and coping?. J Anxiety Disord 2020; 74:102271. DOI: 10.1016/j.janxdis.2020.102271

44. Boyraz G & Legros DN: Coronavirus Disease (COVID-19) and Traumatic Stress: Probable Risk Factors and Correlates of Posttraumatic Stress Disorder. J Loss Trauma 2020; 25(6-7):503–522. DOI: 10.1080/15325024.2020.1763556

45. Roberts AL, Gilman SE, Breslau J, et al: Race/ethnic differences in exposure to traumatic events, development of post-traumatic stress disorder, and treatment-seeking for post-traumatic stress disorder in the United States. Psychol Med 2011; 41(1):71–83. DOI: 10.1017/S0033291710000401

46. Alexander AC, Ali J, McDevitt-Murphy ME, et al. Racial Differences in Posttraumatic Stress Disorder Vulnerability Following Hurricane Katrina Among a Sample of Adult Cigarette Smokers from New Orleans. J Racial Ethn Health Disparities. 2017;4(1):94–103. doi:10.1007/s40615-015-0206-8

47. Holmes SC, Facemire VC, & DaFonseca AM: Expanding criterion a for posttraumatic stress disorder: Considering the deleterious impact of oppression. Traumatol 2016; 22(4):314– 321. DOI: 10.1037/trm0000104

48. Galovski TE, Peterson ZD, Fox-Galalis A: Trajectories of Posttraumatic Stress and Depression in Police and Community Members Following the Violence during Civil Unrest in Ferguson, Missouri. Am J Community Psychol 2018; 62(3-4):433–448. DOI:10.1002/ajcp.12273

49. Moody AT & Lewis, JA: Gendered racial microaggressions and traumatic stress symptoms among Black women. Psychol Women Q 2019; 4(*2*):201–214. DOI: 10.1177/0361684319828288

50. Kokou-Kpolou CK, Megalakaki O, Laimou D, & Kousouri M: Insomnia during COVID-19 pandemic and lockdown: Prevalence, severity, and associated risk factors in French population. Psychiatry Res 2020; 290:113128 DOI: 10.1016/j.psychres.2020.113128

51. Massazza A, Joffe H, Hyland P, Brewin CR: The structure of peritraumatic reactions and their relationship with PTSD among disaster survivors. J Abnorm Psychol 2021;

